# Several Mathematical Aspects on Daily Number of COVID-19 Infection Cases in Eight Southeast Asian Places

**DOI:** 10.1101/2023.07.19.23292810

**Authors:** C.L. Chow, C.H. Cheng, W.K. Chow

## Abstract

The number of daily confirmed infected cases is a key parameter to determine emergency management actions to take. The mathematical characteristics of the daily infection number should be explored for working out appropriate control scheme. Several mathematical aspects on the daily number of infected cases will be discussed in 8 Southeastern Asian places using the confirmed daily infection numbers available in public websites. Phase space diagrams of plotting the daily infection rate estimated numerically against daily infection number on those tests are presented first. Modeling parameters including the Farr’s Law are also discussed. A parameter is proposed to describe the extent of infection by estimating the transient daily infection number divided by the time.

## 1. Introduction

Rapid COVID-19 outbreaks of the fifth wave [1–3] in the Hong Kong Special Administrative Region (HKSAR) started from January 2022. This wave of outbreak is a quick incidence by a few incoming persons carrying the highly infectious Omicron variant infecting over 1.2 million citizens (over 16% of 7 million) in May 2022. The healthcare systems handling testing, transportation, quarantine accommodation, medical treatment and even the temporary dead body storage were all overloaded. More infection cases were reported in August, giving 1.6 million infected in the end of September 2022. The situation appeared smoothened but the number rose again in mid-December with 2.6 million infected up to the end of December 2022 and then calmed down. Many control schemes were removed in early February 2023 except still requiring people to wear mask.

The number of daily confirmed infected cases observed (I(n) at the n^th^ day) is an important parameter in indicating COVID-19 outbreaks. This number should be monitored properly and then estimated for making appropriate decision on emergency management in a city. Locking down the city is an example, which is difficult to implement due to the economic impact. The susceptible number of people to be infected might be reduced with the recovery number increased. The number will also be useful in making decision to relax the control scheme. However, relaxing interflow through free travelling might lead to mass infection by new variant. The public healthcare system might be overloaded again. It is necessary to explore the mathematical characteristics of the daily infection number for working out appropriate control scheme.

In this paper, several mathematical aspects on the number of infected cases I(n) on the nth day will be discussed by taking Hong Kong as an example. The reported confirmed daily infection number available in public websites in seven other places in Asia including Tokyo, Singapore, Shanghai, Beijing, Southern Korea, Taiwan and North Korea [1,4-7] will be compared. The mathematical aspects of the daily infected number in those eight places will be discussed in this paper. The phase space diagram of plotting the daily infection rate estimated against daily infection number on those tests will be presented. These include the variation of I(n) and other patterns such as plotting I(n+1) against I(n) and appropriate fitted equations, phase diagram time rate against I(n), modeling parameters including the Farr’s Law in Hong Kong [8–12].

It is desirable to work out a forecasting parameter based on I(n). As the situation is so complicated, why not going back to a simple approach in just observing the I(n) curve which would result in different emergency management strategy, mitigation, containment or others? A simpler approach monitoring the daily I(n) curve on nth day is more desired, though difficult to achieve. A parameter is needed to indicate the rise or the fall of infection. Tendency of increase in I(n) might probably indicate having new variants inside the places, or outside external infection from countries with high infection number, but adequate mitigation actions. More stringent inspection procedures are needed at entry points. The fall trend of I(n) might lead to relaxing entry inspection and other gathering restriction schemes, if the variants are not so toxic. For example, I(n) appears to be large in the end of December 2022, but the Omicron and new variants are not so toxic. As announced in news, the serious cases might be 1% [13], and causality lower than 0.1% [14]. However, those infected by other variants might be troublesome, and some places like USA advised citizens to wear mask again. Therefore, the daily trends on infection number are still important for taking appropriate action not to overload the public health care systems, or to decide in relaxing control schemes to resume normal living.

## 2. Infection Pattern in Southeast Asia

Different places in SE Asia were infected by the variant Omicron and Delta with the number reported in various sources [1,4-7]. Singapore had many cases first, started on 20 December 2021. They implemented the scheme “live with the virus”. Infection then spread to Tokyo, started from 3 January 2022, and South Korea from 23 January 2022. In Hong Kong, some members under waived scheme did not follow quarantine for 21 days, and spreads out virus, starting on 23 January 2022, infecting over 1.2 million people in 3 months. Omicron and new variants were also spread to China Mainland, Beijing from 19 February 2022, Shanghai from 4 February 2022, Taiwan from 6 March 2022, and North Korea (announced as fever cases, not as COVID-19) from 11 May 2022.

The number of infection cases I(n) on the n^th^ day in those eight places are plotted in Fig. 1a. Macau started to have the daily infection number increased, though that was below 100 a day up to 24 June 2022, with a maximum of 145 on 5 July 2022 [7].

**Fig. 1:**
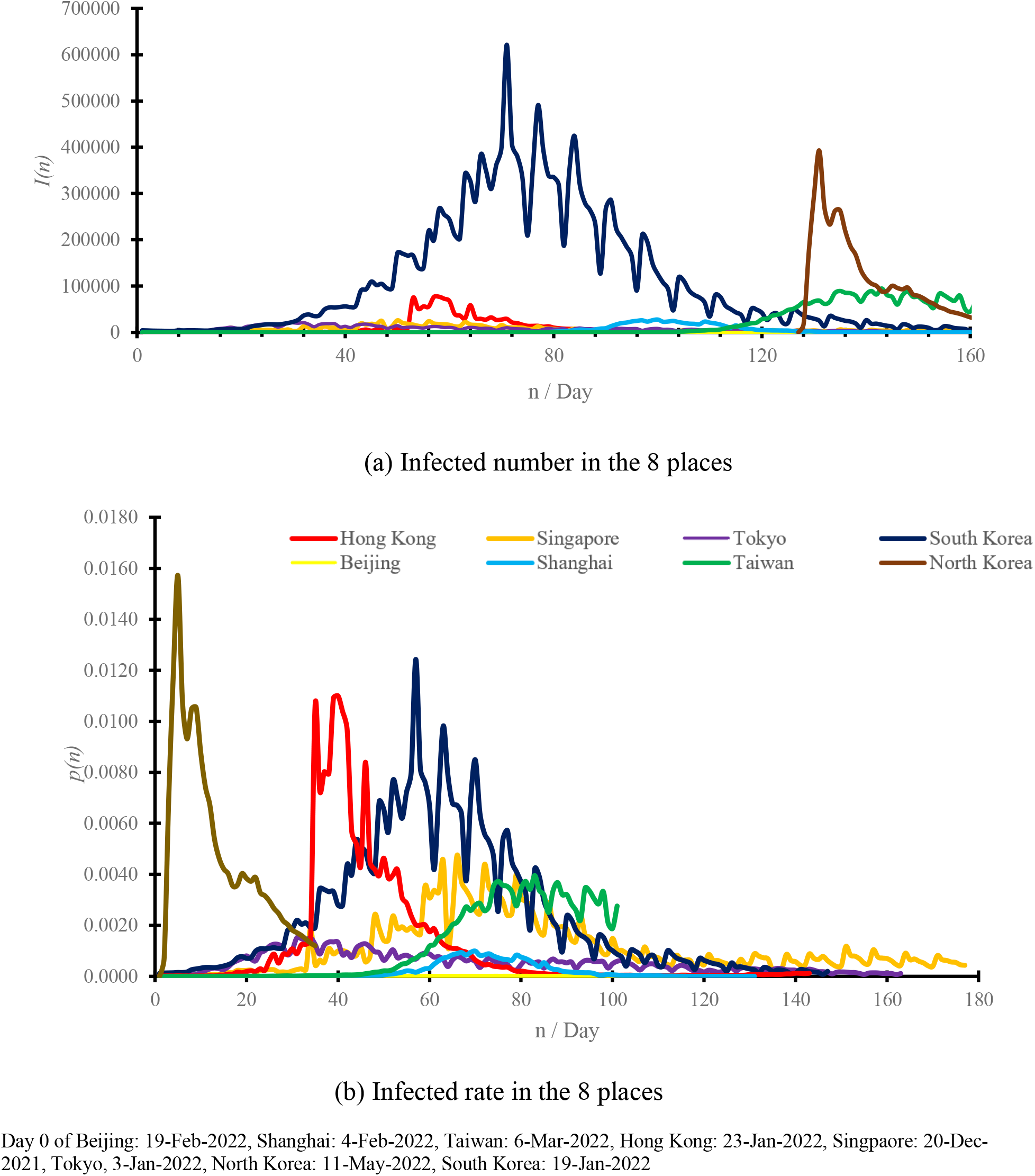
Daily infection number at the 8 places. Data sources: World Health Organization. https://covid19.who.int/ China Briefing. https://www.china-briefing.com/news/china-coronavirus-updates-latest-developments-business-advisory-part-2/ 38 North. https://www.38north.org/2022/05/north-korean-covid-19-fever-data-tracker/ Medriva. https://charts.medriva.com/country/MO

The starting dates at those 8 places are different with different control schemes. Under the very infectious variant Omicron, the number of daily infection cases I(n) rose rapidly in the first few days. I(n) can even go up to a high value as in Fig. 1a, say around 80,000 cases a day in Hong Kong, then drop slowly depending on the control scheme implemented.

The infection ratio p(n) of place is taken as I(n) divided by the total population number N_T_ of the place as:

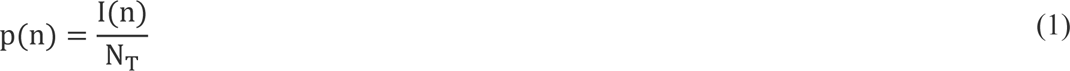

Shifting the starting date of all 8 places to the origin, and plotting p(n) against time (the n^th^ day) gives Fig. 1b.

It can be seen that the curves rose to peak values and then moved down.

Further, the daily infection case I(n+1) on the (n+1)^th^ day is plotted against I(n) in Fig. 2.

**Fig. 2:**
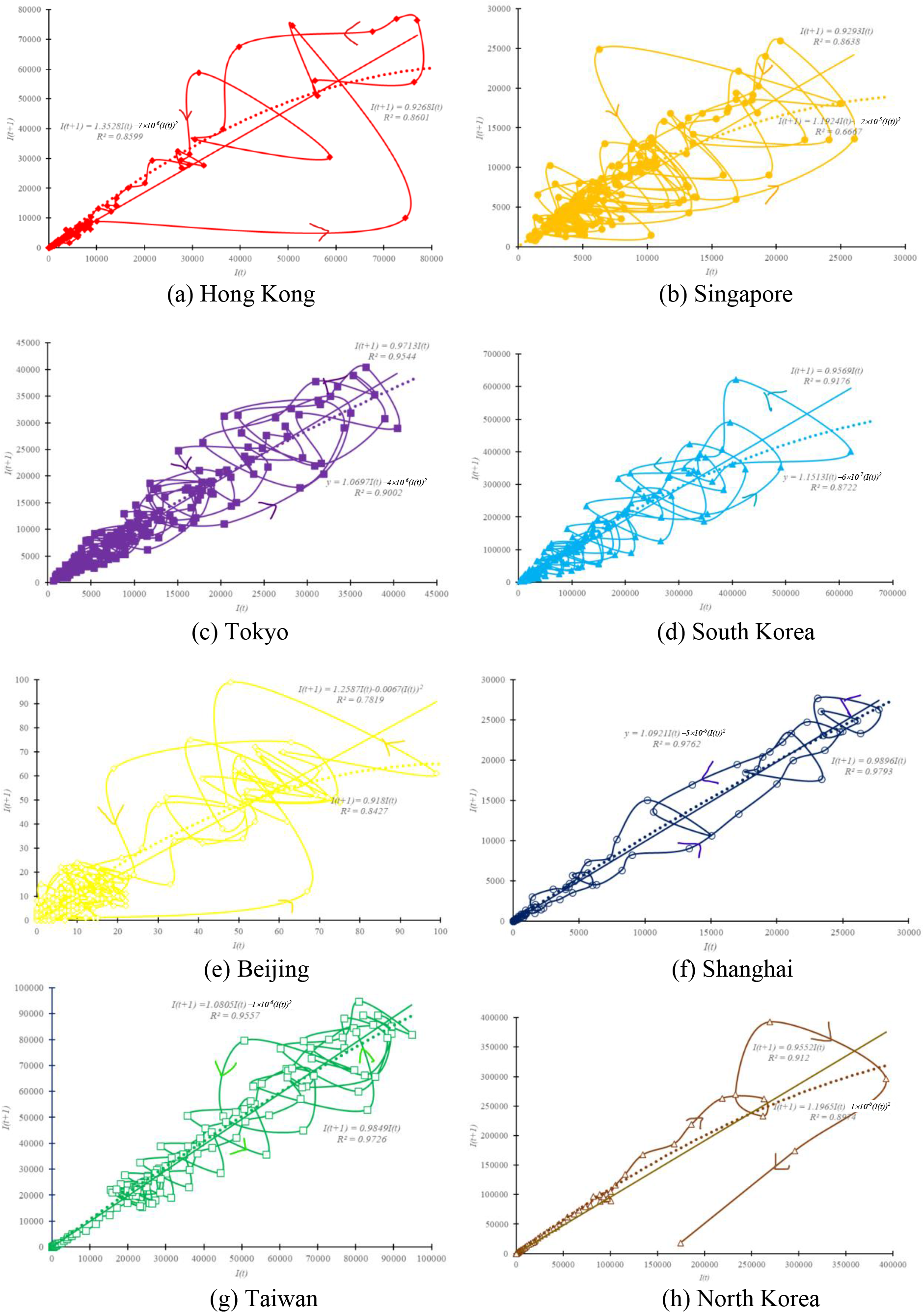
I(n+1) vs I(n) and fitted equations.

Linear equations can be fitted for the 8 places. Hong Kong with correlation coefficient R^2^ of 0.8603 as following:

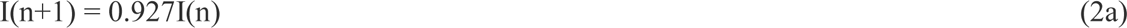

Singapore has correlation coefficient R^2^ of 0.8653:

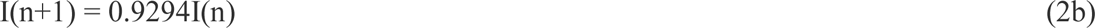

Tokyo has correlation coefficient R^2^ of 0.9544:

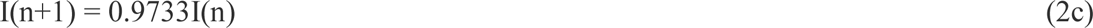

South Korea has correlation coefficient R^2^ of 0.9179:

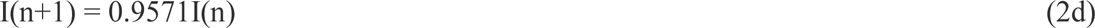

Beijing has correlation coefficient R^2^ of 0.8426:

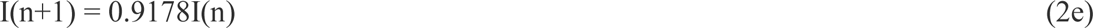

Shanghai has correlation coefficient R^2^ of 0.9793:

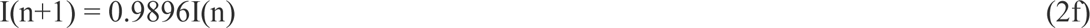

Taiwan has correlation coefficient R^2^ of 0.9727:

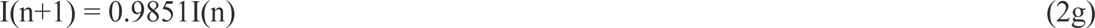

North Korea has correlation coefficient R^2^ of 0.9408:

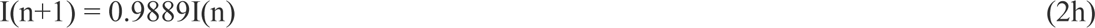

The 8 fitted equations are plotted in Fig. 2. In view of Fig. 2, linear relationship might not be appropriate. Another approach is to describe a more complicated equation in terms of parameters I_o_ and b [15]:

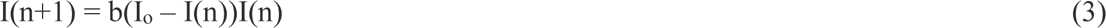

The results for Hong Kong with correlation coefficient R^2^ of 0.8621 are also shown in Fig. 2.

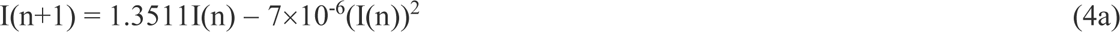

The results for Singapore with correlation coefficient R^2^ of 0.6696 is:

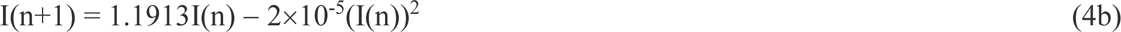

The results for Tokyo with correlation coefficient R^2^ of 0.8995 is:

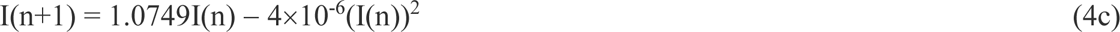

The results for South Korea with correlation coefficient R^2^ of 0.8724 is:

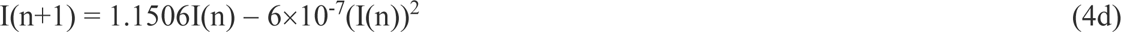

The results for Beijing with correlation coefficient R^2^ of 0.7816 is:

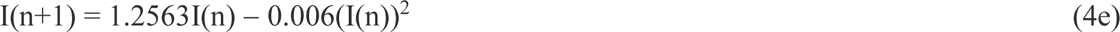

The results for Shanghai with correlation coefficient R^2^ of 0.9762 is:

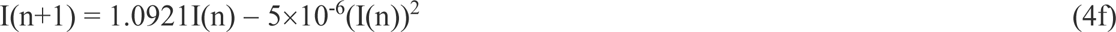

The results for Taiwan with correlation coefficient R^2^ of 0.9556 is:

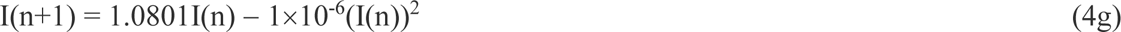

The results for North Korea with correlation coefficient R^2^ of 0.9427 is:

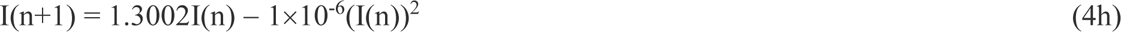

## 3. Phase Space Diagrams

In each place, the daily infection case I gives a rate dI/dn (as time rate is changed with respect to number of days) which can be estimated by numerically using the 5-point method [16]:

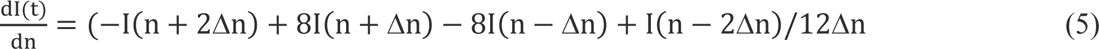

On the n^th^ day, with

Δn = 1 day

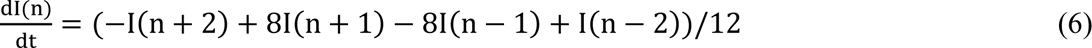

Phase space diagrams showing d(n)/dt against I(n) for all 8 places using equation (6) are plotted in Fig. 3. This gives how the rate changes with I.

**Fig. 3:**
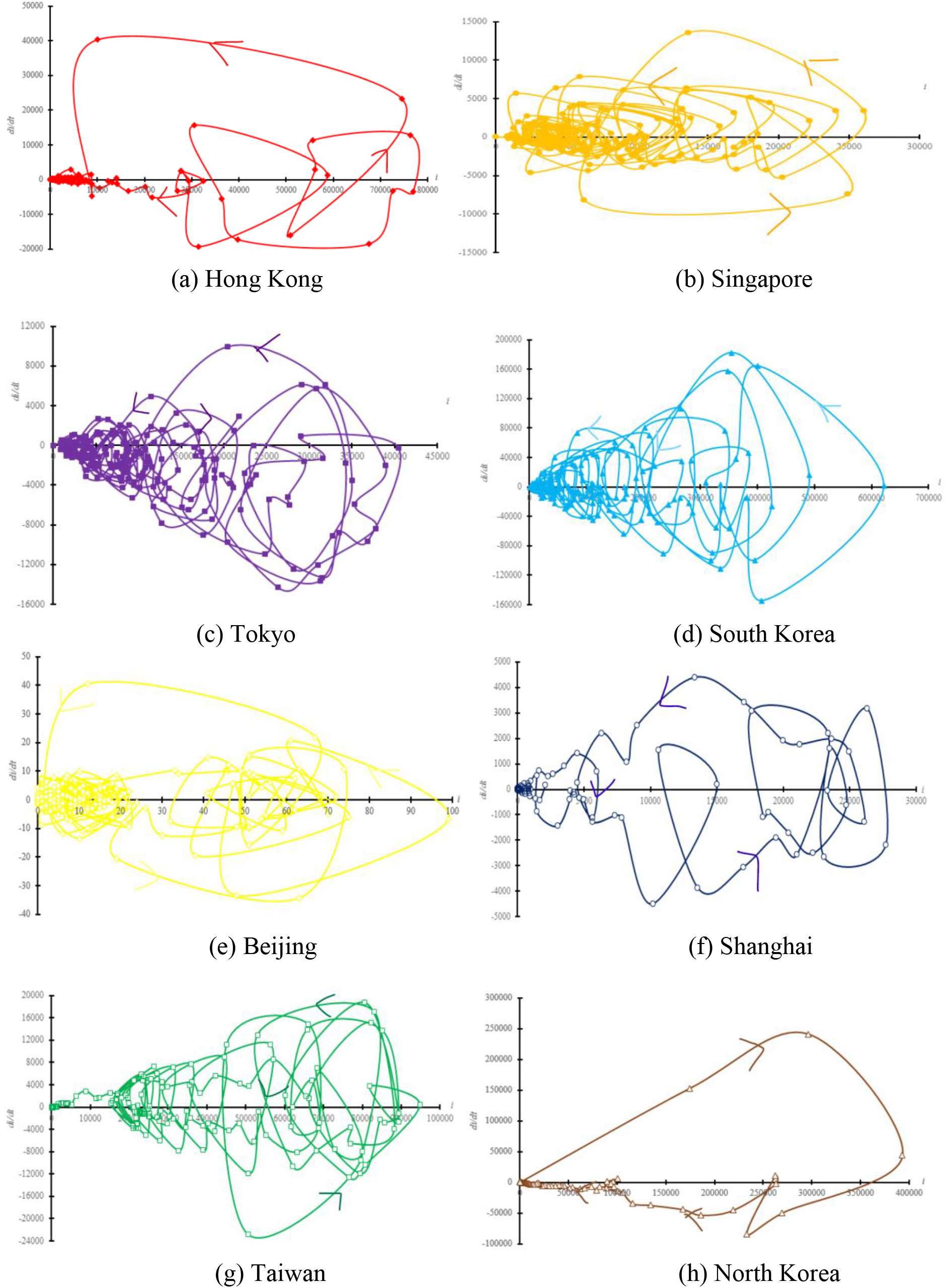
Phase diagram 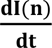 vs I(n)

## 4. Non-linear Dynamics

Early epidemics law by Farr [8–12] was commonly used on COVID-19 outbreak [12] in different Southeast Asian places. Acceleration of new cases and deaths can be predicted by modelling [12] the COVID-19 data.

Two parameters R_1_(n) and R_2_(n) on the n^th^ day are included in Farr’s Law as used in [12] based on prior reports [8,10,11]:

- The first parameter is R_1_(n) on the change of cases or deaths comparing (n+1)^th^ day against the n^th^ day. Subtracting 1 from R_1_(n) would give the percent increase of cases or deaths, R_1_(n) could be understood as the “velocity of spread” [14] of the epidemic. R_1_(n) is the ratio of I(n+1) over I(n):

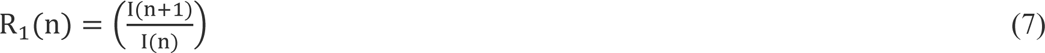
- The second parameter R_2_(n) on day n measures the rate of change of R_1_(n). It compares the R_1_(n+2) against the R_1_(n), interpreted as the acceleration of the epidemic. Dividing the R_1_(n+2) over that before R_1_(n) yields the second ratio R_2_(n) on the n^th^ day.

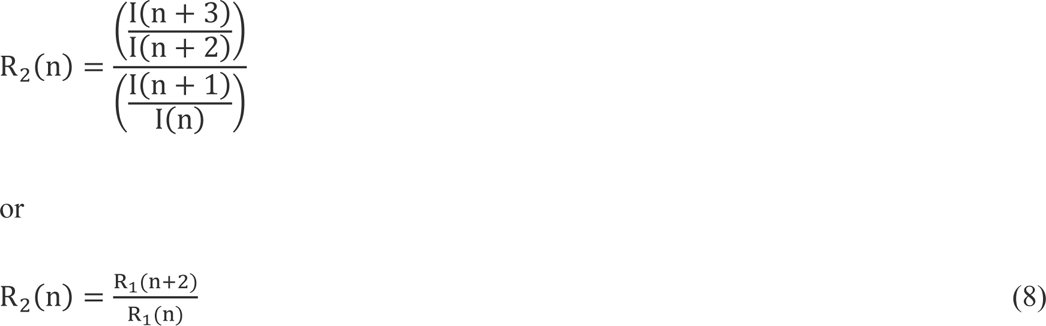

These two numbers can be calculated using epidemic curves and predictive models on I(n) appropriate for the countries concerned. Empirical correlation analysis with available data was studied [8,10,11]. The results suggest that Farr’s law can give an overview of COVID-19 pandemic dynamics.

R_1_ and R_2_ in Hong Kong at critical periods in the first 160 days are plotted in Fig. 4. R_1_ and R_2_ for all 8 places are plotted in Fig. 5.

**Fig. 4:**
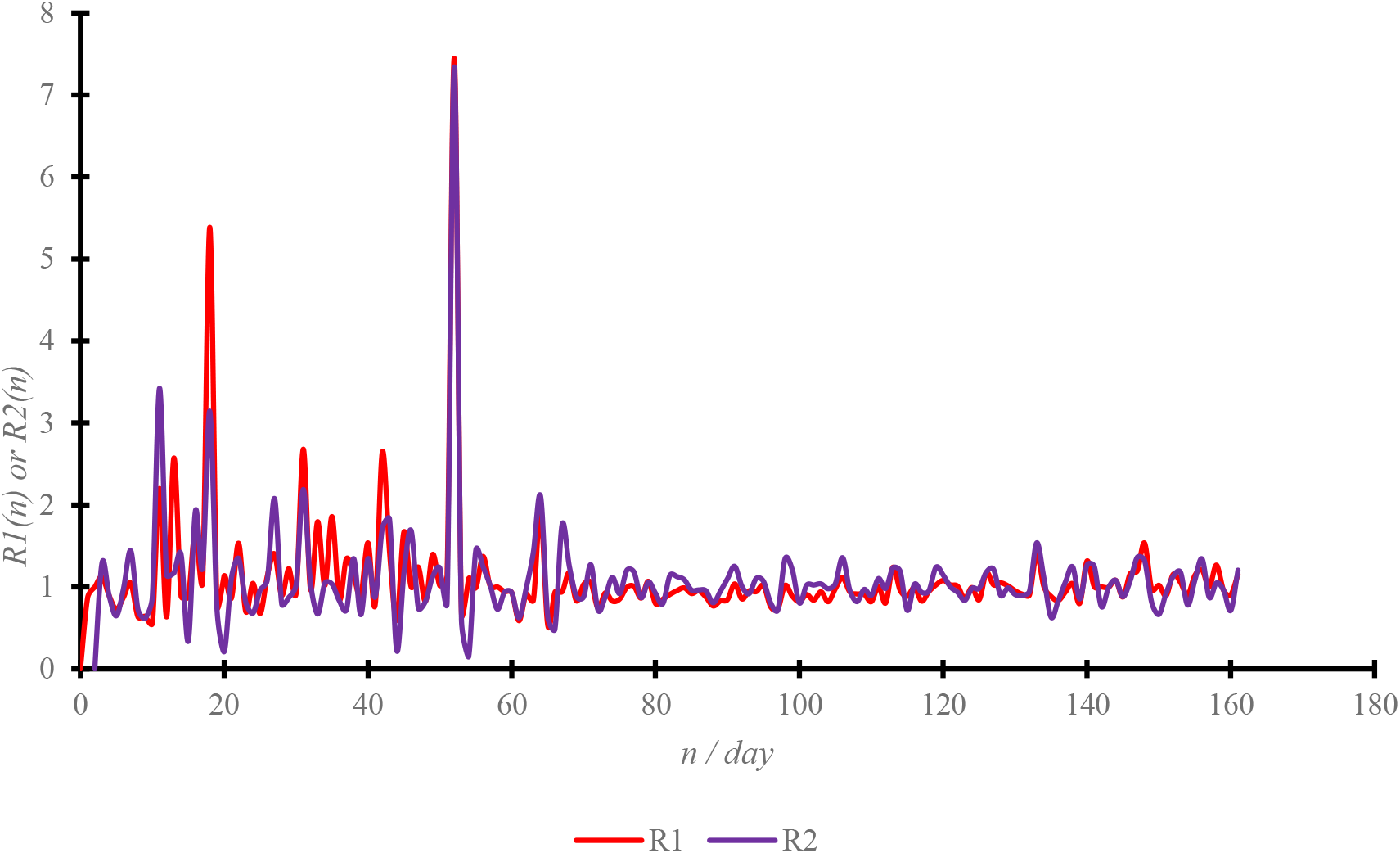
Farr’s Law R_1_ and R_2_ in Hong Kong.

**Fig. 5:**
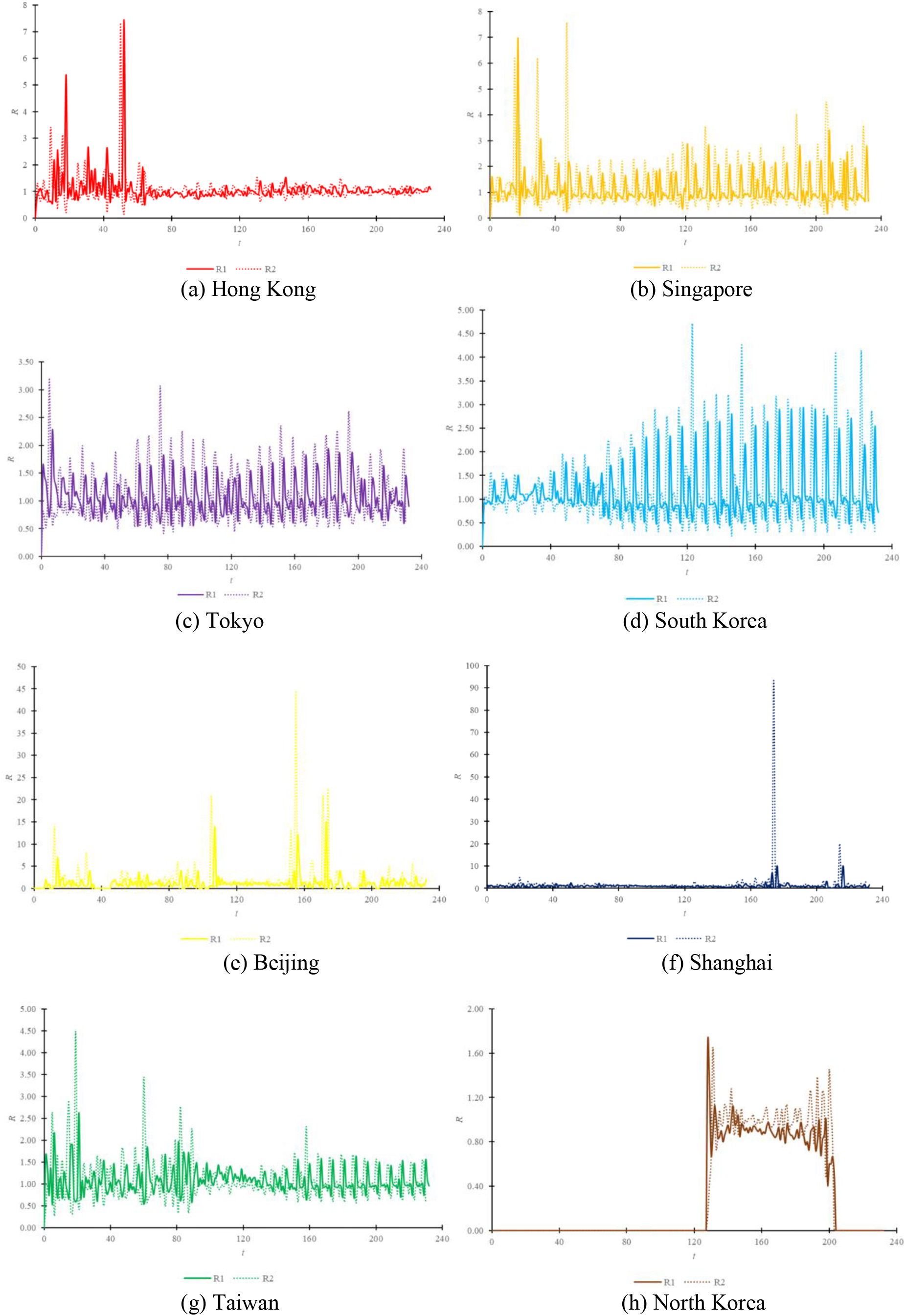
Farr’s Law ratio R_1_/R_2_ for the 8 places.

If taking R_1_ as a constant m:

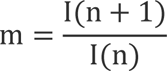

Another graph for I(n+1) against I(n) can be plotted to fit a line to see the value of m:

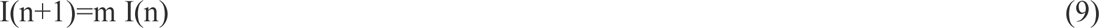

The results are shown in Fig. 2 in the 8 places with equation fitted as in section 2.

Slope m and correlation coefficient were fitted from observed I as in equations (2a) to (2h).

## 5. Monitoring Parameters Proposed

A parameter α on the n^th^ day can be defined to monitor how fast infection increased as:

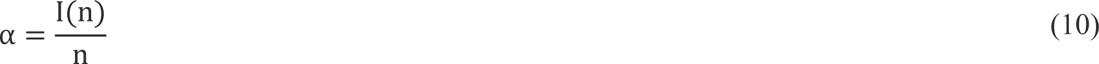

Results of a are plotted against I(n) in Fig. 6.

The following curves are fitted in the 8 places and plotted in Fig. 6:

Hong Kong has correlation coefficient *R^2^* of 0.98:

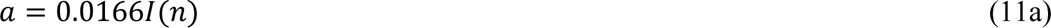

Singapore has correlation coefficient *R^2^* of 0.6086:

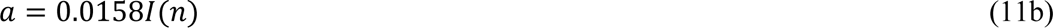

Tokyo has correlation coefficient *R^2^* of 0.4298:

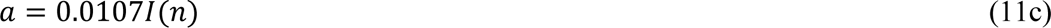

South Korea has correlation coefficient *R^2^* of 0.8986:

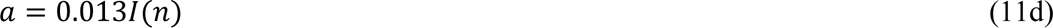

Beijing has correlation coefficient *R^2^* of 0.7079:

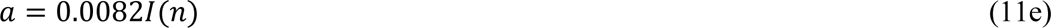

Shanghai has correlation coefficient *R^2^* of 0.9944:

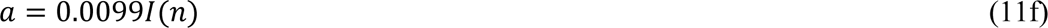

Taiwan has correlation coefficient *R^2^* of 0.9828:

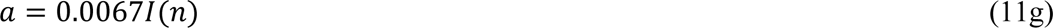

North Korea has correlation coefficient *R^2^* of 0.7362:

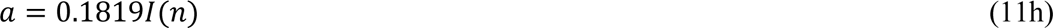

**Fig. 6:**
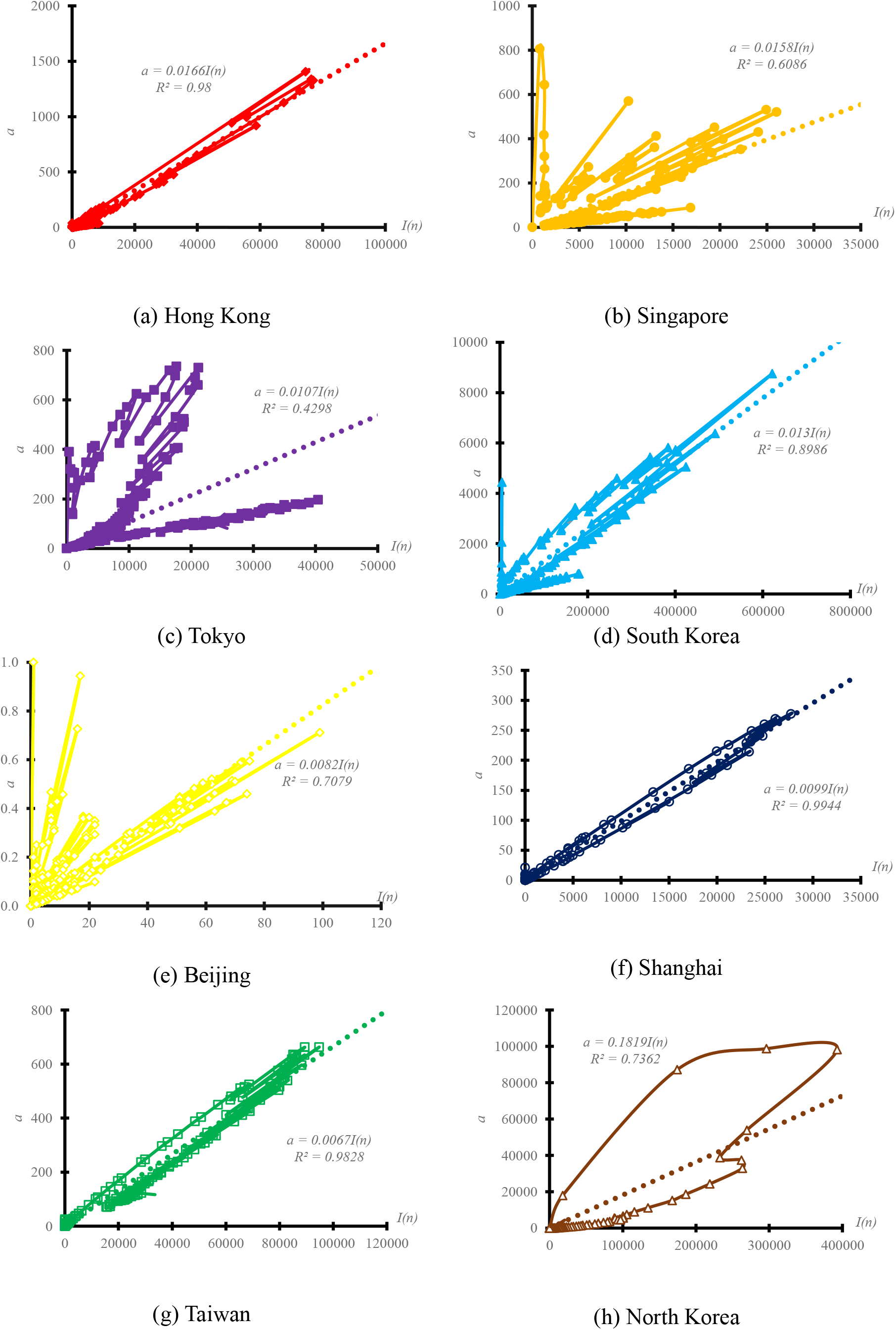
Monitoring parameters a vs I.

Transient results of α with n are also plotted in Fig. 7. In view of Fig. 7, it appears that α_peak_ peak values of α_1_ to 1/3 of α_peak_ takes roughly similar time Δt, around 10 days. That was 13 days for Hong Kong, 4 days for Singapore, 24 days for Tokyo, 4 days for South Korea, 7 days for Beijing, 16 days for Shanghai, 39 days for Taiwan and 6 days for North Korea.

**Fig. 7:**
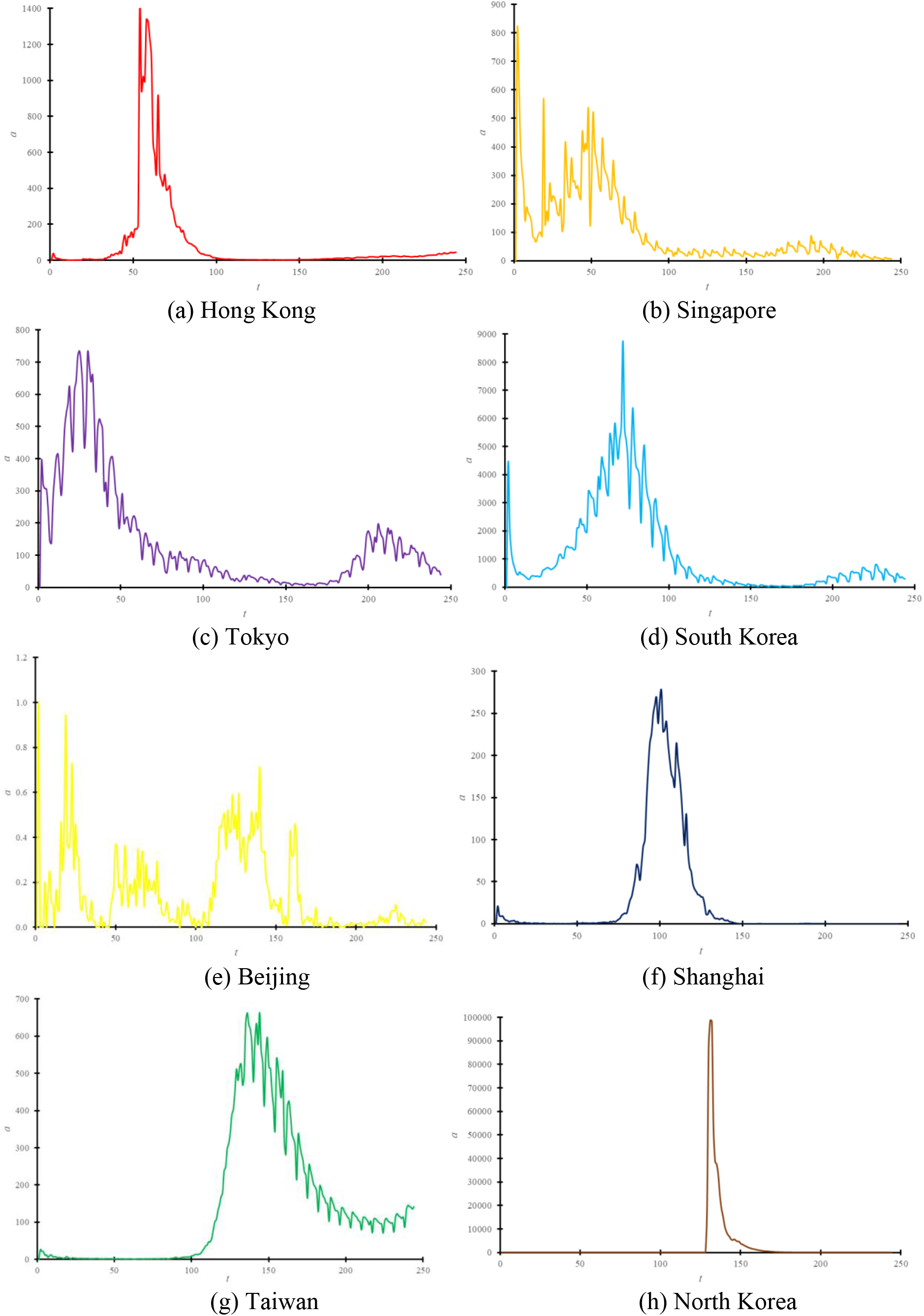
Transient a vs n.

The parameter α is ratio of I(n) divided by n, higher the value, higher the I and shorter the time to get it. But after going through the peak, I drops but n increases, giving smaller α. The value of α dropping to an agreed value, say 30% of peak as in above, can give how long it takes the infection number I to drop. On the other hand, monitoring the increase of α would indicate how fast virus spreads.

Further, control schemes such as tight social distancing, traveling restriction, and limiting services hours of catering services and pubs would give a faster drop of α. On the other hand, allowing opening of those places of concern, but without adequate ventilation provisions of say 6 ACH would give a longer time for α to drop, such as in Tokyo and Taiwan.

The time required to fall by 1/3 in the 8 places in Fig. 8.

**Fig. 8:**
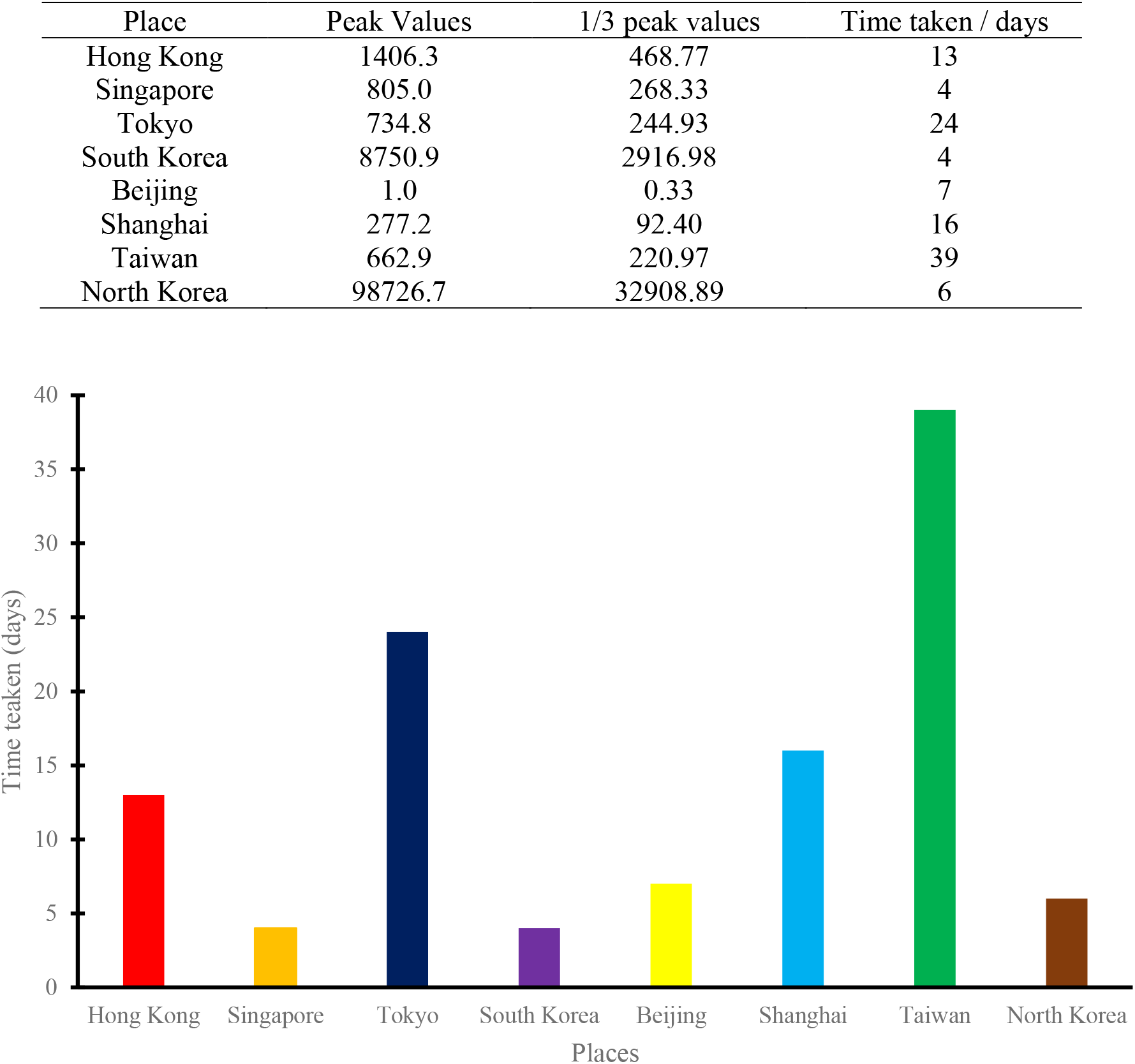
Time taken for α to fall down by one-third peak.

## 6. Rises in August 2022 and then Relaxation

The Omicron BA.5 subvariant is very infectious [17], keeping the number I(n) high in August 2022.

The increase in August in I(n) is shown in Fig. 9.

**Fig. 9:**
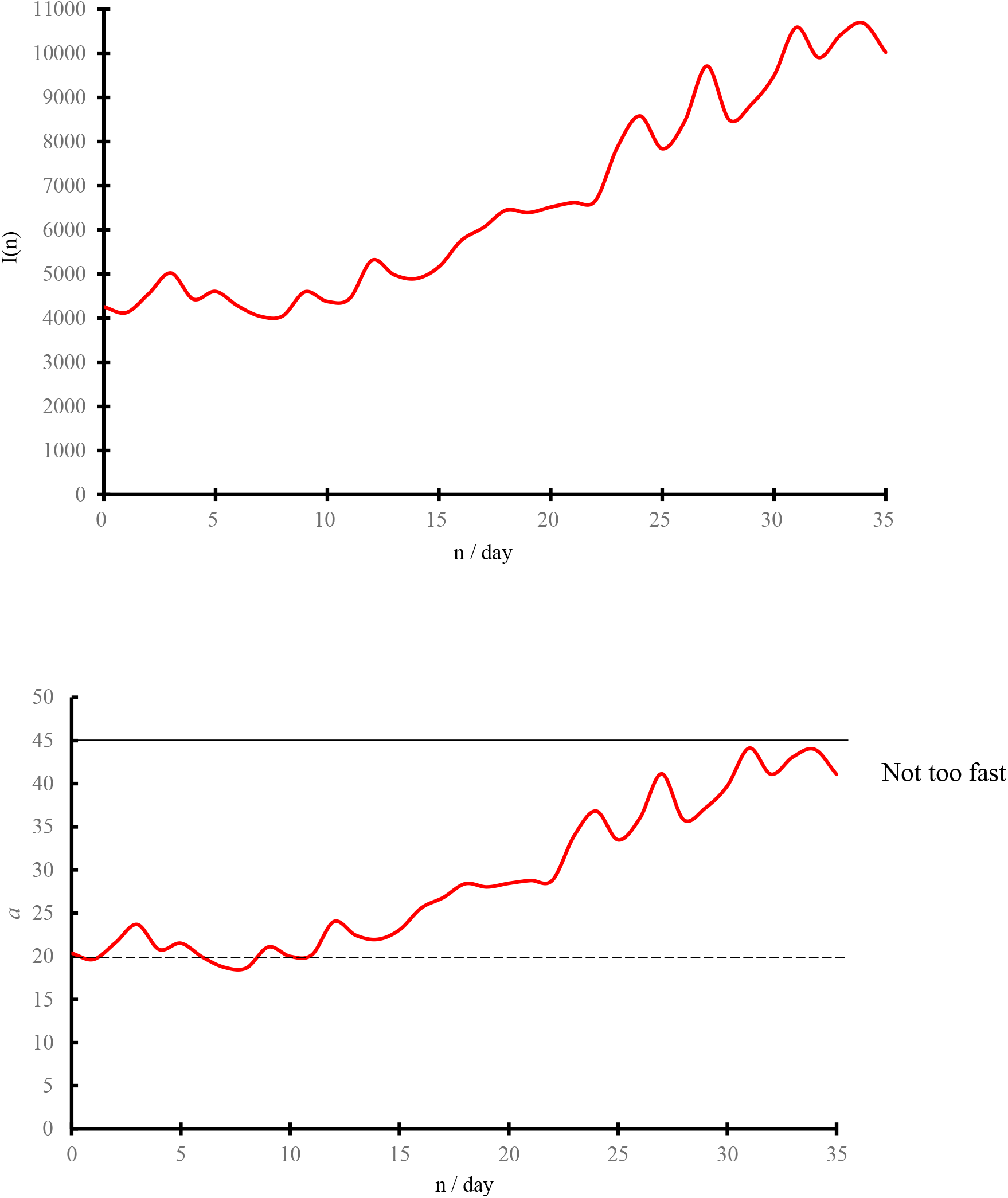
Observation in August 2022.

There are people infected directly, affecting normal life of their colleagues, classmates or family members who might not be infected. Asymptomatic patents might not report to the authority, and the real figures of daily infection might be several times the reported figures.

After analyzing the data, wearing masks is still the best protection. As reported, infection through the following activities are common:

- Maids gathering with inadequate protection, with many unmasked.
- Operation of pre-educational systems of children below 6. Young kids going to school or other tutorial classes.
- Elderly caring services centres or hostels.
- Restaurants without cleaning after serving a table, inadequate ventilation below 6 ACH. Some might even shut down the ventilation to give 0 ACH as caught several times.
- Private parties with many friends.

## 7. Discussion

Effectiveness of COVID-19 strategies adopted by China, Japan, Singapore, and South Korea was analyzed [18] by extracting publicly available data from various official websites, summarizing the strategies implemented in these four countries, and assessing the effectiveness of the prevention and control measures adopted by these countries from October 28, 2020. China, Japan, Singapore, and South Korea implemented different intervention strategies to curb the spread of COVID-19 and maintain lower rates of case-fatality. China, Singapore, and South Korea adopted the containment strategy, while Japan adopted the mitigation strategy. Although Japan’s case-fatality was maintained at a low level, daily new cases increased faster than the other three countries. The study [18] suggested that a mitigation strategy could be inferior to a containment strategy.

The origin of SARS-CoV-2 remains unknown with two leading hypotheses as reported by Sachs et al. 2022 [19]:

- virus emerged as a zoonotic spillover from wildlife or a farm animal, possibly through a wet market.
- virus emerged from a research-related incident, during the field collection of viruses or through a laboratory-associated escape.

## 8. Conclusion

Several mathematical aspects on the daily number of infected cases were discussed in 8 places including Hong Kong, Shanghai, Beijing, Taiwan and Korea later. Confirmed daily infection number available in public websites was used. Phase space diagrams of plotting the daily infection rate estimated numerically against daily infection number on those tests are presented.

## Data Availability

All data produced in the present work are contained in the manuscript.

